# Improving Assessment of Vaccine Effectiveness by Coupling Test-Negative Design Studies with Survival Models

**DOI:** 10.64898/2025.11.30.25341323

**Authors:** Shangchen Song, Matt Hitchings, Yang Yang, Ira M. Longini, the N3C consortium

## Abstract

The test-negative design (TND) has become a widely used observational study design for evaluating vaccine effectiveness, especially during the COVID-19 pandemic. Traditionally, TND has been viewed as a variant of the case-control study and largely limited to use with logistic regression models. In this paper, we first establish that TND can be framed as a special case of a cohort study, thereby opening the door to a wider range of analytical approaches. We then introduce the Prentice, Williams, and Peterson gap-time (PWP-GT) frailty model as a novel method for analyzing TND data, accounting for recurrent infections and time-dependent vaccination status. Through extensive simulation studies, we demonstrate that the proposed model outperforms conventional models commonly applied in TND-based vaccine effectiveness studies. Finally, we apply our method to data from the National COVID Cohort Collaborative, estimating the effectiveness of full and booster doses of Pfizer’s COVID-19 vaccines against both initial infection and reinfection during the Omicron variant circulation period in a real-world setting.

## Introduction

The test-negative design (TND) is an observational study design increasingly used to evaluate vaccine effectiveness, particularly for seasonal influenza ^1^ and more recently during the COVID-19 pandemic ^2,3^. It has been instrumental in assessing post-licensure vaccine effectiveness against SARS-CoV-2 in a real-world setting ^4^. The TND became a preferred approach during the global public health emergency due to its cost-effectiveness, rapid implementation, and ability to be integrated into existing surveillance systems ^5^. Additionally, the TND helps reduce the risk of misclassification bias by confirming the infection status through highly accurate laboratory tests. More importantly, it inherently adjusts for biases related to differential healthcare-seeking behavior between vaccine recipients and non-recipients which are challenging to measure in other study designs ^6–8^.

In the literature ^2,9–11^, the TND is often regarded as a type of case-control study and labeled as a “test-negative case-control study.” Within this framework, investigators typically employ logistic regression as the primary method to compute the adjusted vaccine effectiveness. For example, all 42 TND studies included in a recent meta-analysis ^3^ used either conditional or unconditional logistic regression models. However, relying solely on logistic regression for TND data can present certain limitations. Logistic regression is not well-suited for time-to-event outcomes or addressing dropouts. Furthermore, logistic regression does not provide an ideal framework for modeling recurrent infections or accounting for time-varying vaccination status, both of which are common features of infectious disease outbreaks. Previous work including Sullivan et al. (2016) ^6^, Jackson et al. (2013) ^7^, and Sullivan et al. (2023) ^12^ has highlighted directly or indirectly that the TND may be cohort-like within the subpopulation of symptomatic, tested care-seekers. Shi et al. (2023) ^8^ discussed the potential to use time-to-event models for analyzing TND data, based on some theoretical justification for application of such models to case-control studies ^13^.

In this work, we first demonstrate that the TND can be treated as a special case of a cohort study, allowing for the application of analytic methods beyond logistic regression. We then introduce the Prentice, Williams, and Peterson gap-time (PWP-GT) frailty model and its applicability to the analysis of TND data, especially when recurrent infections are possible. Previous studies ^14–16^ indicate that prior infection history or hybrid immunity (prior infection plus vaccine) provide more protection than vaccine alone for SARS-CoV-2 variants. Similarly, for influenza, vaccine immunogenicity was improved by prior infection than prior vaccination ^17–19^. These findings motivated our consideration of recurrent infections when estimating vaccine effectiveness. Next, we conduct a series of simulations to compare the performance of the proposed method against existing models used in TND studies. Finally, we apply the proposed method to the National COVID Cohort Collaborative datasets to estimate vaccine effectiveness in a real-world setting.

## Methods

### Background of TND

A TND study usually begins by identifying a population (often geographically defined) where the pathogen of interest is actively circulating and some individuals have received the vaccine under study. At the testing facilities within this population, as illustrated in Figure 1, patients presenting relevant clinical symptoms are recruited if they are tested for the pathogen of interest. These patients are then divided into two groups: those who test positive for the pathogen are categorized as “cases”, while those who test negative become the “controls”. The symptoms of these controls are assumed to be attributable to other co-circulating pathogens on which the vaccine has no effect.

**Figure 1.**
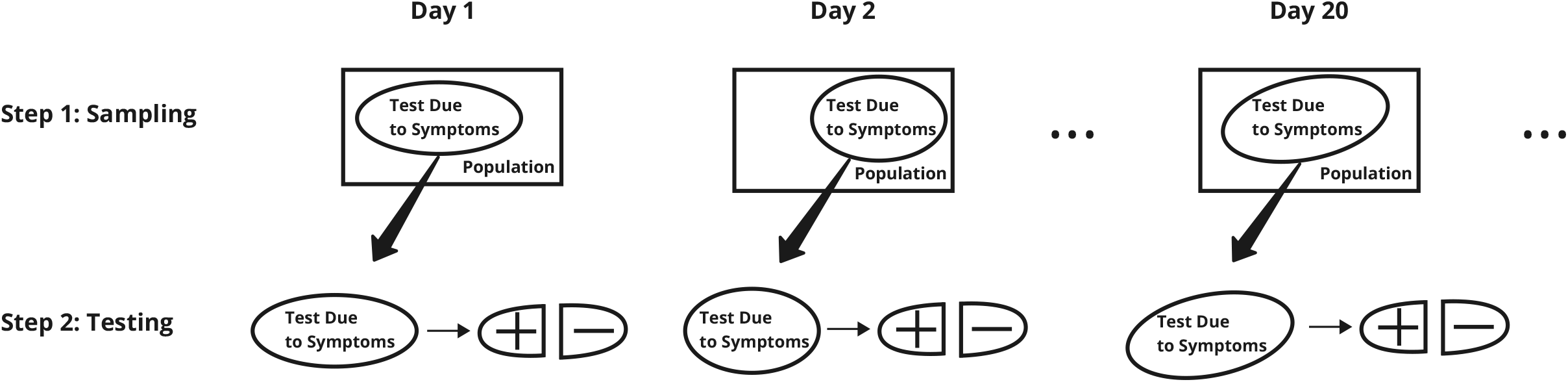
Sampling and testing of the TND. In a population, the testing due to target symptoms acts like a lasso to capture the individuals over time. The infection status is ascertained only after an individual is recruited.

The vaccine effectiveness is subsequently calculated using the formula Vaccine Effectiveness = 1 − Odds Ratio of vaccination between cases and controls. This odds ratio is usually adjusted for key confounders using unconditional logistic regression. Alternatively, if there are enough cases and controls, matching can be applied to reduce confounding by pairing or stratifying based on similar characteristics. In this situation, conditional logistic regression is used, which is closely related to the Cox regression model when matching is based on infection time ^20^. The validity of TND relies on two important assumptions: (1) among individuals who present with the clinical syndrome eligible for enrollment, the probability of being tested for the target pathogen does not depend on which pathogen actually caused the symptoms; and (2) the vaccine of interest has no biological effect or social effect on nuisance pathogens causing similar symptoms as the pathogen of interest, where the social effect means that people vaccinated against the target pathogen are more likely to be vaccinated against nuisance pathogens. We denote infection status by *I*, symptom status by *s*, vaccination status by *V*, and testing by *T*. Here symptom status refers to the presence of clinical symptoms that may lead to medical attention. We use subscript as a shorthand for the value of each symbol, e.g., *V*_0_ (*V* = 0) and *V*_1_ (*V* = 1) indicating unvaccinated and vaccinated, respectively. Unlike other symbols, *I*_0_ here should be interpreted as infection by a nuisance pathogen, instead of no infection at all. In a simple TND setting where the outcome is binary and confounders are suppressed, the vaccine effectiveness is estimated by

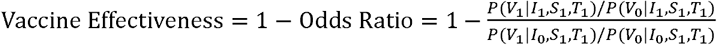

Following the natural data-generating process of vaccination, infection, symptom onset, and testing, we can expand the Odds Ratio as

Odds

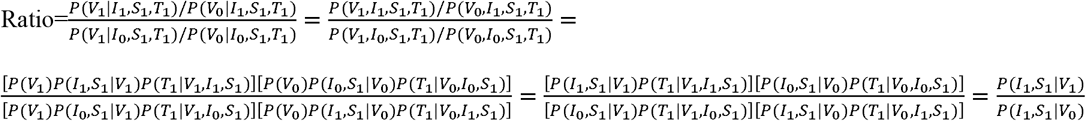

where the last equation follows from *P*(*T*_1_|*V*_1_, *I*_1_, *s*_1_) = *P*(*T*_1_|*V*_1_, *I*_0_, *s*_1_) and *P*(*T*_1_|*V*_0_, *I*_1_, *s*_1_) = *P*(*T*_1_|*V*_0_, *I*_0_, *s*_1_) by assumption (1) and *P*(*I*_0_, *s*_1_|*V*_0_) = *P*(*I*_0_, *s*_1_|*V*_1_) by assumption (2). It should be noted that there is no need to assume that the probability of testing is not affected by vaccination status. Interestingly, the odds ratio in the TND turns into a relative risk in the population sense, which meets the standard definition of Vaccine Effectiveness =1-Relative Risk. It is also clear that vaccine effectiveness estimates from TND studies should be interpreted as vaccine effectiveness against medically-attended symptomatic infections, which is particularly valuable as it reflects the burden on healthcare systems ^21,22^.

### Rethinking the TND study: From case-control-like to cohort-like

The fundamental difference between case-control and cohort studies lies in the sampling of subjects. In cohort studies, participants are selected based on exposure (e.g., vaccination status in vaccine studies) or randomly from the population. In contrast, case-control studies select participants based on their outcome (e.g., infection status). The data distribution in cohort studies is represented as either *P*(*I*|*V*) or *P*(*I,V*), while in case-control studies it is *P*(*V*|*I*).

Although TND studies are often viewed as case-control studies, they more closely resemble cohort studies. In a TND study, the outcome (infection status) is not used for recruitment but is determined only after participants are enrolled. This differentiates it from typical case-control studies as well as nested case-control studies (unless post-hoc subsampling or matching is performed). In TND studies, participants are randomly sampled from individuals who seek testing for a pathogen due to relevant symptoms, representing a conditioned subset of the population. The data distribution in TND studies is represented by *P*(*I*|*V,s*_1_, *T*_1_) or *P*(*V,I*|*s*_1_, *T*_1_). Therefore, a TND study can be viewed as a conditioned cohort study, or alternatively, as a regular cohort study within a subpopulation. This argument can be further supported by re-writing the above odds ratio as

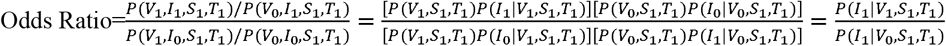

where we utilized assumption (2) again, *P*(*I*_0_|*V*_0_, *s*_1_, *T*_1_) = *P*(*I*_0_|*V*_1_, *s*_1_, *T*_1_). If *P*(*I*_1_|*V*_1_, *s*_1_, *T*_1_) and *P*(*I*_1_|*V*_0_, *s*_1_, *T*_1_) imitate population level probabilities, we can analyze the TND data as a conditional cohort, using statistical models commonly used for cohort studies such as survival models. This is equivalent to requiring *P*(*I*_1_, *V*_1_|*s*_1_, *T*_1_) and *P*(*I*_1_, *V*_0_|*s*_1_, *T*_1_), and ultimately *P*(*I*_1_|*s*_1_, *T*_1_) and *P*(*I*_0_|*s*_1_, *T*_1_), to be similar to the population level proportions. Clearly, only TND studies that used all or a random sample of tested patients satisfy this requirement, whereas those matching each test-positive patient with a prespecified number of test-negative ones do not.

### Relationship among the TND, the cohort study design, and the nested case-control study design

In nested case-control studies, cases and at-risk controls are sampled at each case time from a predefined cohort to form the at-risk set for that time point. Conditional logistic regression or Cox regression may then be used to estimate odds or hazard ratios associated with risk factors such as vaccination. If the sampling weights are known, the baseline hazard can also be reliably estimated. If the sampling probability is 1, then the nested case-control is equivalent to the cohort study design. A typical TND is a special (or conditional) cohort study restricted to the subpopulation who seek healthcare and get tested if they become ill during the study period. Enrollment occurs at care□seeking and testing regardless of the potential test result, and infection status is ascertained after enrollment. If a subset of test-negative controls is sampled to measure exposure history, referred to as a case□density (time□matched) subsampling scheme, then the TND is not a special cohort study anymore but still a special nested case-control study (here, we focus on TND where exposure history is measured on all enrolled subjects, not the case□density subsampling scheme). Viewing TND as a conditional cohort allows us to fit survival models to estimate both hazard ratios and baseline hazards, while naturally accounting for (i) time□varying vaccination, (ii) recurrent infections via event□order strata, and (iii) community-level calendar□time hazards. Estimation of baseline hazards is especially important when the model contains frailties in order to reflect heterogeneity in the baseline hazard across individuals or clusters (Therneau et al., 2003) ^23^.

### PWP-GT Frailty Model

We proposed the Prentice, Williams, and Peterson gap-time (PWP-GT) frailty model, an extension of the Prentice, Williams, and Peterson model ^24^ and the frailty model ^25^ based on Cox regression ^26^, to directly account for recurrent infections and time-dependent vaccination status in TND studies. In comparison, recurrent infections are modeled as binary outcomes in logistic regression, with previous infections and vaccination status treated as indicator covariates at pre-specified time points, resulting in loss of information ^27^.

The PWP-GT frailty model handles the recurrent infections by assuming that a subject is not at risk for the *s*^th^ infection until they have experienced the (*s* − 1)^th^ infection. Data associated with different infections are assigned into separate strata, each corresponding to a distinct infection. Therefore, this model provides more flexibility in allowing distinct baseline hazard and coefficients of covariates for each infection. The PWP-GT frailty model further assumes that the baseline hazard depends on the elapsed time since the previous infection—hence the term “gap-time”—rather than the total time from initial risk exposure, accounting for host-level risk changes such as individual immunity due to sequential infections. To complement host-level risk factors, community-level risk covariates can be incorporated into the model by adjustment, such as case numbers across different calendar time periods. Because multiple infections in a subject introduce significant within-subject correlation, the PWP-GT frailty model incorporates a frailty term to explicitly account for this dependency. Alternatively, a robust sandwich variance estimator ^28^ can be used instead of a frailty term to handle within-subject correlations, using a variance adjustment similar to generalized estimating equations models ^29^. When recurrent events are sparse, the frailty term can also be adapted to represent other correlation structures, such as the location of testing facilities. Finally, time-dependent covariates are used to model vaccination status, allowing the model to capture dynamic changes due to different doses or vaccination regimens.

Suppose a pathogen can cause up to s infections in a subject. The PWP-GT frailty model has the following hazard function *λ*_*si*_ (*t*) for the *i*^th^ subject against the *s*^th^ infection.

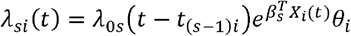

where *λ*_0*s*_ (*t* − *t*_(*s*−1)*i*_) is the baseline hazard for the *s*^th^ infection at time *t* − *t*_(*s*−1)*i*_ since the (*s* − 1)^th^ infection occurred to subject *i*. This is a shared term across all subjects who have experienced the (*s* − 1)^th^ infections. *X*_*i*_ (*t*) is a vector of covariates for the *i*^th^ subject at time *t*, which may include both time-dependent and time-independent variables affecting the hazard of the *s*^th^ infection, and *β*_*s*_ is the associated vector of coefficients. *θ*_*i*_ is the frailty term, assumed to follow a Gamma distribution *θ*_*i*_ ∼ Gamma(*α, β*) with a mean of 1. The estimation and inference of the model parameters can be achieved by the penalized partial likelihood approach, which is implemented in standard statistical software packages, e.g., the *coxme* R package ^30^. The vaccine effectiveness estimate for *s*^th^ infection is given by Vaccine Effectiveness_*s*_ = 1 − exp (*β*_*s*_) if *X*_*i*_ (*t*) indicates the vaccination status.

### Simulation Studies

We conducted a series of hazard-based simulation studies to evaluate the performance of our proposed models compared to other commonly used vaccine effectiveness estimation models in TND studies. We also examined the performance of the proposed models in a cohort study where healthcare-seeking behavior is not adequately controlled for, illustrating potential biases in vaccine effectiveness estimates. We considered situations where vaccine effectiveness remains constant over time and where it progressively wanes in a piecewise manner across short, medium, and long-term periods, with a specific focus on the performance of the PWP-GT frailty model under the second situation (progressive waning of vaccine effectiveness), which is more realistic for SARS-CoV-2 vaccines during the COVID-19 pandemic.

We investigated seven scenarios in the first situation (constant vaccine effectiveness), defined by combinations of study designs and analytic models. The first two scenarios applied the PWP-GT model in a TND study, incorporating either a frailty term (Scenario 1, PGF-TND) or a robust sandwich variance estimator (Scenario 2, PGR-TND). The third scenario used conditional logistic regression in a TND study (Scenario 3, CL-TND). The fourth and fifth scenarios used unconditional logistic regression in a TND study, with adjustment for calendar time (Scenario 4, ULA-TND) or without such adjustment (Scenario 5, UL-TND). Finally, the sixth and seventh scenarios applied the PWP-GT model in a cohort study, again using a frailty term (Scenario 6, PGF-Cohort) or a robust sandwich variance estimator (Scenario 7, PGR-Cohort).

We simulated 600 cohorts, each containing 30,000 participants. For each cohort, we first conducted a cohort study where all participants were followed longitudinally, with their vaccination and infection status tracked irrespective of whether they sought testing. Using the same cohort, we then simulated a TND study nested in the cohort, focusing only on individuals who sought testing due to symptoms caused by COVID-19 or other co-circulating respiratory pathogens. Healthcare-seeking behavior, which influences both vaccination and infection, was simulated as an unmeasured variable that could not be included in the model. This introduced confounding bias in the cohort study; in the TND study, this bias could be controlled by conditioning on testing. eFigure 1 includes a directed acyclic graph illustrating this setup. Furthermore, our simulations only consider symptomatic infections, as TND studies primarily capture symptomatic cases.

In each simulation, a subset of the individuals who received a full dose of vaccine further received a booster dose. Although a full course includes two partial doses separated by several weeks for most SARS-CoV-2 vaccines, for simplicity we did not consider the time gap between the partial doses in the simulation. The vaccine effectiveness against initial infection was set at 50% for full doses and 60% for the booster dose, while for reinfection,Vaccine effectiveness was set at 70% and 80%, respectively. A frailty term, following a Gamma(5, 0.2) distribution, was incorporated into the simulation for infection times. Detailed procedures for data generation and model implementation are provided in eAppendix 1 to 2. The simulation codes are available on the GitHub repository at https://github.com/songuf/TND-Survival.

The simulation results across 600 cohorts are presented in Figure 2 including the average estimates of the four vaccine effectiveness and their quantile intervals (2.5%, 97.5%) for each of the seven scenarios outlined above. The probability of the 95% confidence interval (CI) covering the true vaccine effectiveness was shown beside each quantile interval. Our analysis revealed that the PWP-GT frailty model in a TND study (Scenario 1, PGF-TND) achieves both minimal empirical bias relative to the true vaccine effectiveness and the best CI coverage at the nominal level. The PWP-GT model with a robust sandwich variance estimator in a TND study (Scenario 2, PGR-TND) performed slightly worse than the PWP-GT frailty model in a TND study (Scenario 1, PGF-TND), especially for the initial infection, suggesting that the robust sandwich estimator may not sufficiently address the within-subject correlation. The conditional logistic regression in a TND study (Scenario 3, CL-TND) showed more biases in the vaccine effectiveness estimates and substantially worse CI coverage than two PWP-GT models in a TND study (Scenario 1 and 2, PGF-TND and PGR-TND). The performance of the unconditional logistic regression in a TND study with adjustment for calendar time (Scenario 4, ULA-TND) was similar to that of the conditional logistic regression in a TND study (Scenario 3, CL-TND) for the full doses but slightly inferior for the booster dose. The unconditional logistic regression in a TND without adjustment for calendar time (Scenario 5, UL-TND) was associated with the most severe biases and nearly zero CI coverage probabilities, likely because the unconditional odds ratio does not approximate the hazard ratio in the absence of adjustment for calendar time. In the context of cohort studies, both the PWP-GT models (Scenario 6 and 7, PGF-Cohort and PGR-Cohort) yield inflated vaccine effectiveness estimates and poor CI coverage probabilities, likely due to their failure to control for the confounding effect of healthcare-seeking behavior. eAppendix 4 provides more detailed performance metrics.

**Figure 2.**
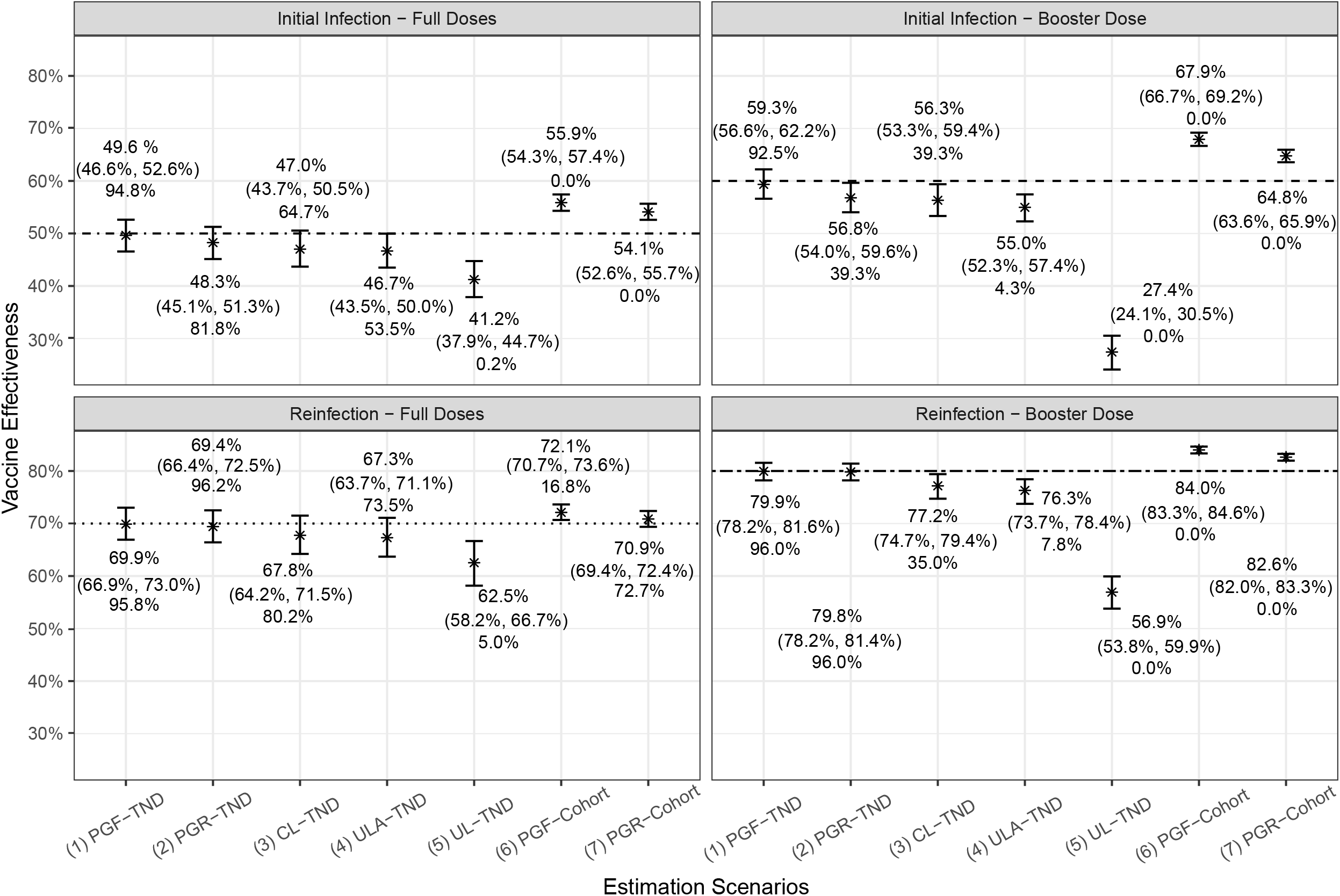
Simulation results when vaccine effectiveness remains constant over time: average vaccine effectiveness estimates [asterisks, top row of text label] with (2.5%, 97.5%) quantile intervals [vertical bars, mid row of text label] and 95% CI coverage probabilities [bottom row of text label] across 600 cohorts for the seven scenarios of study design and used model.

We also simulated TND studies with vaccine effectiveness that wanes over time as piecewise functions to assess the proposed PWP-GT frailty model under more realistic conditions. We considered solely the full doses against the initial infection and reinfection for simplicity. The vaccine effectiveness was set to decline over every 3-month period, with 50%, 40%, and 30% for the short, medium, and long term, respectively, against the initial infection and 80%, 70%, and 60% for the short, medium, and long term, respectively, against the reinfection. Additional details are available in eAppendix 3.

The simulation results for waning VE are shown in Figure 3. The average simulation estimates are very close to the true vaccine effectiveness. The increasing uncertainty of the vaccine effectiveness estimates over time, particularly against the initial infection, is expected as the number of infection events associated with long-term vaccination are relatively lower due to censoring of subsequent vaccination, infection and the end of study. The 95% CI coverage probabilities are close to the nominal level. eAppendix 5 provides more detailed performance metrics. These results reveal that the proposed model successfully captures and estimates the waning trend of the vaccine effectiveness.

**Figure 3.**
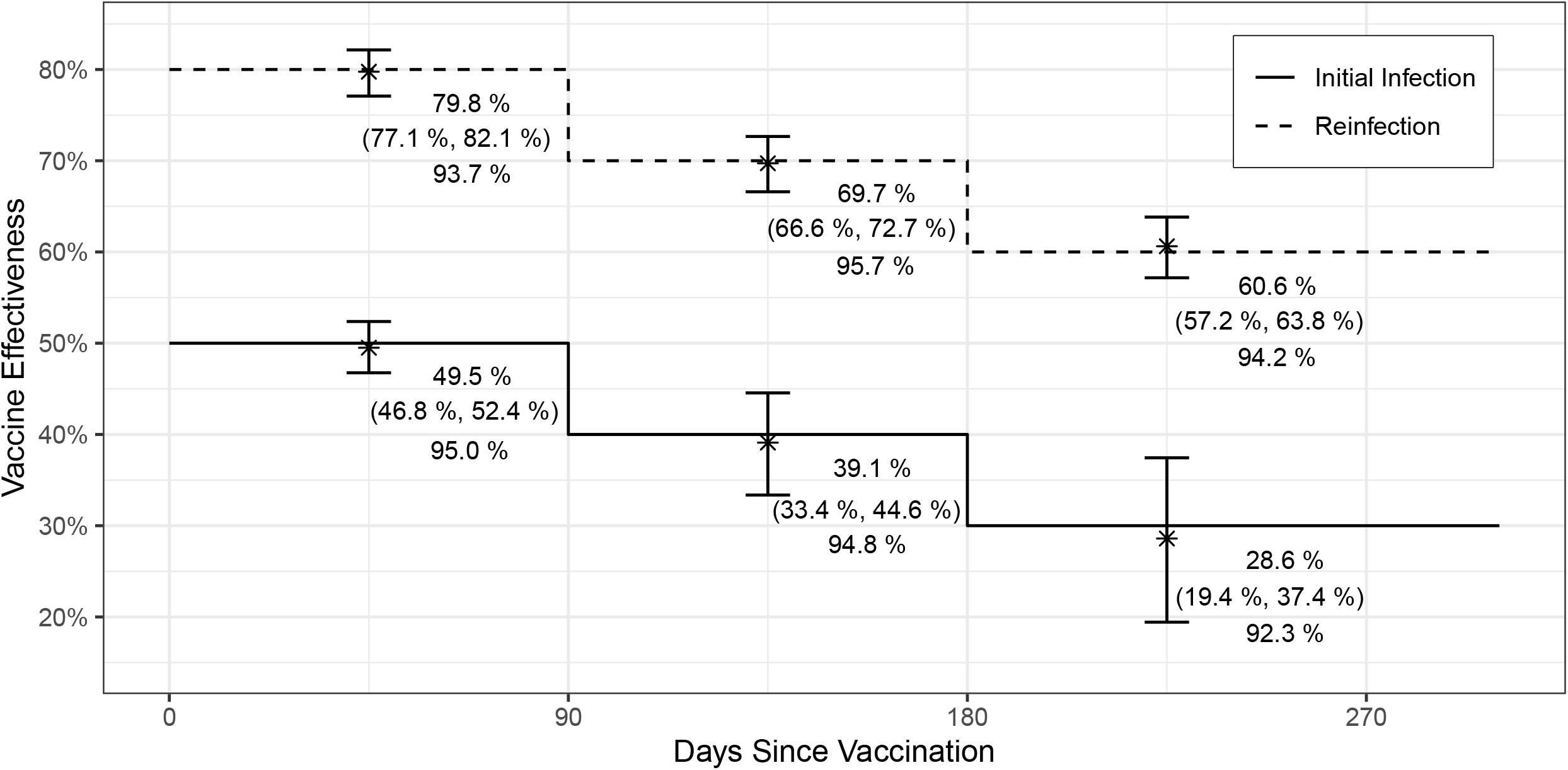
Simulation results when vaccine effectiveness wanes over time: average vaccine effectiveness estimates [asterisks, top row of text label] with (2.5%, 97.5%) quantile intervals [vertical bars, mid row of text label] and 95% CI coverage probabilities [bottom row of text label] across 600 cohorts of the PWP-GT frailty model.

In summary, the proposed PWP-GT frailty model demonstrated satisfactory performance for both constant vaccine effectiveness and waning vaccine effectiveness. Compared to conventional vaccine effectiveness estimation methods, this model achieves smaller bias and better 95% CI coverage via proper handling of recurrent infections and time-varying vaccine status.

### National COVID Cohort Collaborative Real Data Analysis

We applied the proposed PWP-GT frailty model to the National COVID Cohort Collaborative datasets ^31^. The National COVID Cohort Collaborative platform, developed as a centralized repository, aggregates electronic health record data of COVID-19 testing records from healthcare centers across the United States. It represents one of the most extensive and reliable multicenter databases for COVID-19 testing records in real-world settings. More details about the National COVID Cohort Collaborative platform are provided in eAppendix 7.

This analysis focused on estimating the vaccine effectiveness of the Pfizer-BioNTech COVID-19 mRNA vaccine (BNT162b2) against Omicron-related initial infection and reinfection of SARS-CoV-2. As the National COVID Cohort Collaborative dataset does not uniformly specify infection variants, Omicron-related infections were identified by considering infection events in the dataset from December 1, 2021, when the first U.S. case attributed to the Omicron variant was reported, up to April 02, 2024. The vaccine effectiveness was evaluated for both the short-term (14-180 days) and long-term (greater than 180 days) periods following the administration of the full dose (2 doses) and the full dose plus the first booster dose (3 doses).

Following the data processing steps (eAppendix 8), the final dataset consists of 212,887 subjects eligible for inclusion in this analysis. All subjects received the full doses of the BNT162b2 vaccine, with 78,998 (37%) receiving the booster dose. In terms of infection status of the Omicron variants, 155,449 individuals were never infected, 56,204 had only the initial infection, and 1,208 experienced reinfections. The distribution plots of age, location (state), infection time, and vaccination time in the final dataset are provided in eAppendix 9.

In the PWP-GT frailty model, we adjusted for gender, age group, race, and pre-Omicron infection status as time-independent covariates. To account for the risk of infection, we divided it into host-level and community-level components. The host-level risk of infection is captured by individual covariates and the gap-time scale in the model, which is independent of calendar time and determined solely by the individual (e.g., internal immunity). To represent the community-level risk of infection (e.g., community spread), we included time-specific case numbers (updated every four months) as a time-dependent covariate. Additionally, we incorporated a frailty term at the state level instead of subject level to account for within-location correlation, recognizing that reinfection is relatively rare. eAppendix 8 gives more detailed information about the model set-up. The analysis codes and outputs are available upon request within the National COVID Cohort Collaborative Enclave (https://covid.cd2h.org/enclave/) under Project ID RP-7FEF96.

Table 1 presents the estimated vaccine effectiveness of the full doses and the booster dose against Omicron-related initial infection and reinfection, evaluated at 14-180 days and beyond 180 days post-vaccination. The table also includes the associated 95% CI and standard errors of the vaccine effectiveness estimates. For the initial Omicron-related infection, the full doses of the BNT162b2 vaccine demonstrate a vaccine effectiveness of 25.80% within 14-180 days following vaccination, which decreased moderately to 19.03% after 180 days. The booster dose was associated with a vaccine effectiveness of 52.11% within 180 days of boosting, but its effectiveness declined sharply to 9.88% after six months. This substantial drop is likely due to the long time since vaccination of individuals within the 180+ days group (average ∼400 days). Regarding the Omicron-related reinfection, the vaccine effectiveness estimate of the full doses was 73.48% within the six-month vaccination window, with a slight decline to 68.71% beyond 180 days. The vaccine effectiveness estimates of the booster dose against reinfection are comparable to those of the full doses, 72.94% for the first 14-180 days and 65.05% six months or more since the booster dose. No significant violation of the proportional hazards assumption was detected in the graphs of the scaled Schoenfeld residuals.

**Table 1.**
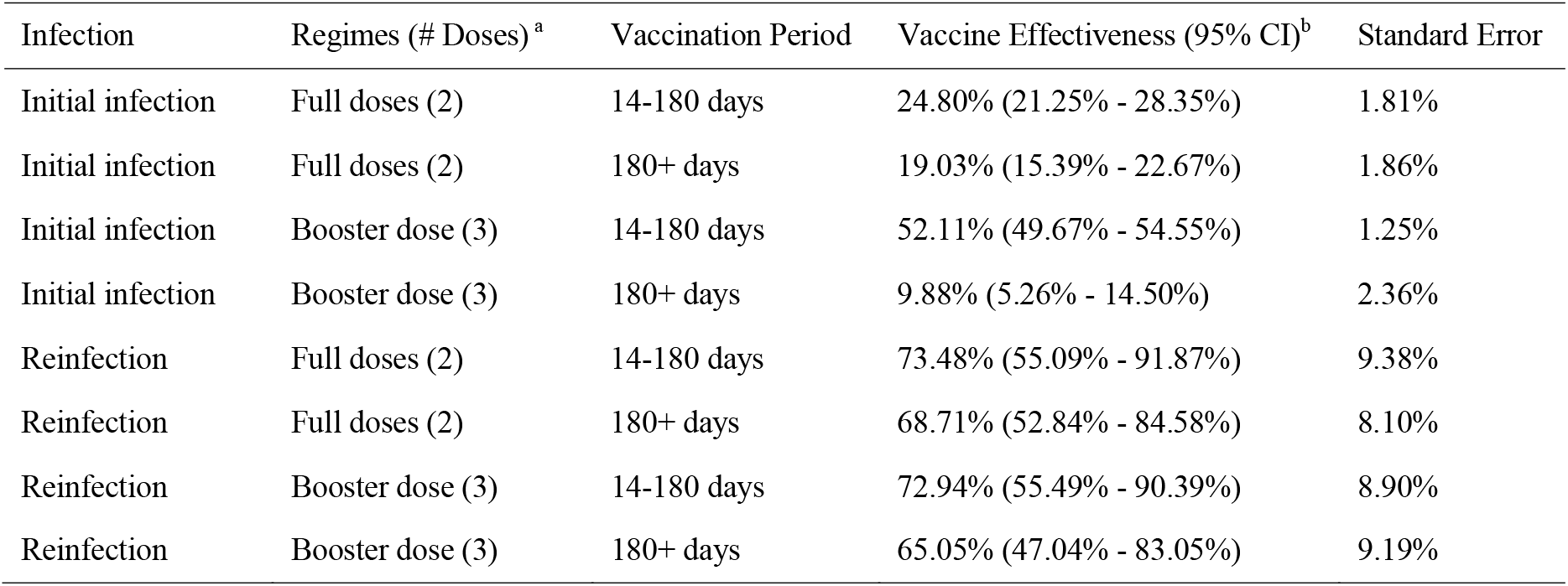
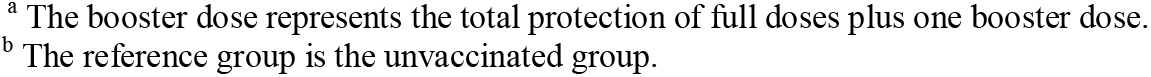
Vaccine Effectiveness Estimation Results, National COVID Cohort Collaborative Data, N= 212,887.

| Infection | Regimes (# Doses)<sup>a</sup> | Vaccination Period | Vaccine Effectiveness (95% CI)<sup>b</sup> | Standard Error |
| --- | --- | --- | --- | --- |
| Initial infection | Full doses (2) | 14-180 days | 24.80% (21.25% - 28.35%) | 1.81% |
| Initial infection | Full doses (2) | 180+ days | 19.03% (15.39% - 22.67%) | 1.86% |
| Initial infection | Booster dose (3) | 14-180 days | 52.11% (49.67% - 54.55%) | 1.25% |
| Initial infection | Booster dose (3) | 180+ days | 9.88% (5.26% - 14.50%) | 2.36% |
| Reinfection | Full doses (2) | 14-180 days | 73.48% (55.09% - 91.87%) | 9.38% |
| Reinfection | Full doses (2) | 180+ days | 68.71% (52.84% - 84.58%) | 8.10% |
| Reinfection | Booster dose (3) | 14-180 days | 72.94% (55.49% - 90.39%) | 8.90% |
| Reinfection | Booster dose (3) | 180+ days | 65.05% (47.04% - 83.05%) | 9.19% |

<sup>a</sup> The booster dose represents the total protection of full doses plus one booster dose. <sup>b</sup> The reference group is the unvaccinated group.

These estimates are similar to those in the literature ^11,32–34^ and comprehensive meta-analyses ^3^. Our results show that in real-world settings, 3 doses of the BNT162b2 vaccine were able to provide protection against Omicron-related medically-attended symptomatic infections for up to six months, and both the full doses and the booster dose offered high levels of short- and long-term protection against subsequent infections among people had experienced an initial Omicron-related infection.

## Discussion

Unlike traditional case-control studies or other outcome-driven sampling design frameworks, TND studies randomly sample individuals presenting clinical symptoms who are to be tested for a pathogen, making them more analogous to cohort studies within a subpopulation. In this study, we proposed a novel use of the PWP-GT frailty model in TND studies to address recurrent time-to-event outcomes, individual-level heterogeneity, and time-dependent vaccination status. Our simulation studies demonstrated that the proposed model yields less biased vaccine effectiveness estimates and better nominal coverage probability of confidence intervals, compared to traditional vaccine effectiveness estimation models commonly used in TND studies. Furthermore, we applied the model to the National COVID Cohort Collaborative dataset for estimating real-world effectiveness of full and booster doses of Pfizer’s COVID-19 mRNA vaccine against subsequent infections, and our results are in line with existing estimates.

The TND remains a relatively new study design, and its unique structure makes it susceptible to various sources of confounding and biases that require careful consideration^8,35–39^. This work introduces survival models as a powerful alternative to traditional logistic regression approaches, offering significant advantages in addressing recurrent infections and dynamic vaccination history. The survival models better capture the dynamics of both infection risks and vaccine status (a time-dependent variable) at the individual-level and thus outperform the logistic regression models. For the logistic regression models (CL-TND, ULA-TND and UL-TND), an individual contributes to the likelihood or conditional likelihood only for the times he/she was tested. In contrast, in the survival models, an individual contributes to the partial likelihood for each event time regardless of whether they were tested at that time or not, as long as they were considered at risk at that time point (i.e., before a positive test of the target event). Additionally, the frailties in the PWP-GT models better capture the correlation of events within each individual, whereas the logistic models treat the two infections as independent events, which could be another reason why the PWP-GT models outperform logistic regression. Consistent with other literature ^40^, our simulation results also underscore the risk of severe bias when using logistic regression in TND studies unless calendar time is appropriately handled.

The adoption of survival analysis allows TND studies to capture nuanced temporal dynamics of infection and immunity, including time-varying infection hazards and waning vaccine effectiveness, which are increasingly critical for optimizing vaccination strategies and other public health interventions. Survival approaches also facilitate meta-analysis of vaccine effectiveness across multiple types of studies. Traditionally, it is challenging to combine vaccine effectiveness estimates based on hazard ratios from cohort studies and those based on odds ratios from TND studies^3^. Although odds ratios approximate relative risk in many situations, the relative risk does not necessarily approximate the hazard ratio (HR) unless the baseline cumulative hazard (Λ_0_) is small, i.e., 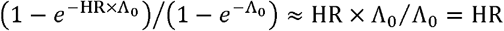 holds only if Λ_0_ is small, which may not be always true. In addition, for conditional logistic regression to produce an odds ratio close to the hazard ratio, the at-risk set at each case time should be formed as in a Cox regression, i.e., each enrolled person should enter the at-risk set at all case times up to their own infection (positive test) or censoring (last negative test). Because this is not necessarily a routine practice in most TND studies, pooling hazard ratio- and odds ratio-based vaccine effectiveness estimates requires either common effect scales (e.g., re-estimating TND with a hazard model) or separate meta-analyses for different effect-measure types. The capacity of TND studies to contribute hazard-based estimates will greatly enrich the data sources for more reliable vaccine effectiveness meta-analysis.

In this work, we adopted the classic TND in which all eligible and enrolled individuals were included in the analyses. If a subset of controls is to be sampled or matched to the case at each event time, the TND essentially becomes a nested case-control design. Conditional logistic regression has been widely applied to nested case-control studies, where the estimated odds ratios are equivalent to the hazard ratios ^41^. However, such equivalence does not hold in the presence of individual-level frailties, as these frailties cannot be canceled in the partial likelihood.

There are at least two potential extensions of our models for broader applications. First,Vaccine effectiveness could be modeled as varying continuously over time. While a piecewise approach is generally sufficient to capture the waning effectiveness in most cases, a continuous-time framework provides a more complete picture at a finer resolution. This can be achieved using time-varying coefficients, e.g., through smoothing splines, or employing Bayesian methods incorporating external information from literature on the temporal pattern of immune responses. Second, for certain infectious diseases where the risk of infection is strongly influenced by the cumulative hazard over the total time rather than the interval between two consecutive events, the gap-time scale could be replaced by the total-time scale. This adjustment may better reflect how immunity jumps and wanes in response to multiple infections or immunization events.

Despite these promising results, our work has several limitations and practical guidance. First, as an inherent limitation of the TND, the method is based on a subpopulation of individuals seeking medical care, and extending these vaccine effectiveness estimates from TND studies to the general population requires additional data sources or assumptions. For example, to obtain vaccine effectiveness against infection, we need the probability of seeking medical attention given infection. Second, similar to Cox regression models applied to observational studies in general, causal interpretation cannot be reached for the vaccine effectiveness estimated by the proposed model within TND studies, as unmeasured confounders are difficult to control for in the absence of randomization, and Cox hazard ratios are not causally interpretable^42,43^. Third, we did not consider the impact of possible prior infection by earlier variants before the Omicron era in the data analysis. We also relied on electronic health record to identify Omicron infection histories, and so may have been unable to include a certain number of asymptomatic and subclinical infections that did not lead to medical visits. These undetected infections could have altered the population immunity landscape and hence affected the vaccine effectiveness estimates. However, it has been well documented that the effectiveness of BNT162b2 against any infection (regardless of symptoms) is relatively low;^44–46^ if undetected infections were equally likely among people with different vaccination histories, and the resulting impact on our estimates of vaccine effectiveness against detected infections may be minor. Moreover, our PWP-GT frailty framework is specifically designed for scenarios where reinfections are fairly frequent. When recurrent infections are rare, for example, a single epidemic season of influenza ^47,48^, the proposed framework reduces to the regular Cox regression with frailties. Finally, our assumption of no social effect of the vaccine of interest on nuisance pathogens may be overly strong in some situations, e.g., elderly people vaccinated against the SARS-CoV-2 are probably more likely to be vaccinated against influenza, and healthcare workers are often required to be vaccinated against multiple pathogens.

In summary, the application of advanced survival models to TND studies provides more reliable estimates of vaccine effectiveness under real-world conditions and thereby better informs public health decision-making. We call for methodological research and data collection regarding potential confounders that affect behaviors related to exposure,Vaccination, healthcare-seeking, and testing to improve causal interpretation of vaccine effectiveness estimates based on TND studies.

## Supporting information

eAppendix

## Data Availability

Simulation codes are available on the GitHub repository at https://github.com/songuf/TND-Survival. Access to the N3C data analysis codes and data is provided through the N3C Data Enclave for users who have an approved protocol and a data use request from an institutional review board. Additional details on accessing the N3C data is provided on https://covid.cd2h.org/enclave/.

## Ethics Approval

This study was deemed exempt by the University of Florida institutional review board (IRB202301849), which was accepted by N3C Data Access Committee (DUR-RP-7FEF96). The N3C data transfer to NCATS is performed under a Johns Hopkins University Reliance Protocol # IRB00249128 or individual site agreements with NIH. The N3C Data Enclave is managed under the authority of the NIH; information can be found at https://ncats.nih.gov/n3c/resources.

## Acknowledgement

Authorship was determined using ICMJE recommendations.

The real data analysis described in this publication were conducted with data or tools accessed through the NCATS N3C Data Enclave https://covid.cd2h.org and N3C Attribution & Publication Policy v 1.2-2020-08-25b supported by NCATS U24 TR002306. This research was possible because of the patients whose information is included within the data and the organizations (https://ncats.nih.gov/n3c/resources/data-contribution/data-transfer-agreement-signatories) and scientists who have contributed to the on-going development of this community resource [https://doi.org/10.1093/jamia/ocaa196]. Further details are described in eAppendix 10.

