## Supplementary material for "Improving Assessment of Vaccine Effectiveness by Coupling Test-Negative Design Studies with Survival Models": eAppendix

### 1 R Functions Used for VE Models in Simulation Studies

| Model | Study | R Function |
| --- | --- | --- |
| PGF | TND | *coxme::coxme* |
| PGR | TND | *survival::coxph* |
| CL | TND | *survival::clogit* |
| ULA | TND | *stats::glm* |
| UL | TND | *stats::glm* |
| PGF | Cohort | *coxme::coxme* |
| PGR | Cohort | *survival::coxph* |

eTable 1 R Functions Used for VE Models in Simulation Studies

### 2 Simulation Settings – Vaccine Effectiveness (VE) Remains Constant over Time

The data generation for simulation studies where VE remains constant over time were based on the Directed Acyclic Graph (DAG) in eFigure 1 and consisted of two steps:

- **First Step:** We simulated data on confounders ($C$), Healthcare-seeking behavior [HSB] ($H$), and vaccination status ($V_{t}$ ), considering these variables didn’t depend on other variables.
- **Second Step:** We then simulated variables of infection status ($I_{t}$), symptom presentation ($S$), and testing status ($D$).

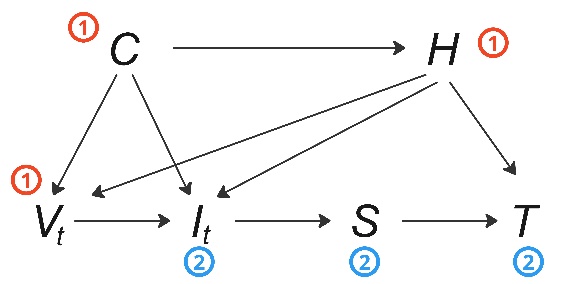

eFigure 1 Diagram of variable simulation. Initially, variables circled by red 1, vaccination ($V_{t}$), confounders ($C$), and HSB ($H$), were simulated. Subsequently, infection ($I_{t}$), symptoms ($S$), and testing ($T$) circled by blue 2 were simulated.

**First Step: Simulation of 𝐶, 𝐻, and** $\boldsymbol{V}_{\boldsymbol{t}}$

For potential confounding factors ($C$), three time-independent characteristic variables, age group ($X_{1}$, $X_{2}$), gender ($X_{3}$), and risk group ($X_{4}$), were incorporated. Age group was differentiated as: adults (60%) for $\left( X_{1},X_{2} \right)=(0, 0)$, children (20%) for $\left( X_{1},X_{2} \right)=(1, 0)$, and elderly (20%) for $\left( X_{1},X_{2} \right)=(0, 1)$. Gender allocation was balanced with female (50%) $X_{3}=0$ and male (50%) $X_{3}=1$. The high-risk population was represented as $X_{4}=0$ and set at 10%.

The simulation of the HSB ($H$) was achieved by setting the logit of the probability of $H=1$, correlating to the sum of a linear function of variables in $C$ and an independent error term from a standard normal distribution to account for unmeasured confounders:

$logit\left( P\left( H=1 | C \right) \right)=3+1.5X_{1}+2X_{2}-X_{3}-3X_{4}+0.5Z$, where $Z\sim N(0,1)$

For vaccination status, we simulated full doses (2-dose primary series, simplified as one uptake) and a booster dose (3rd dose), referring to the latter simply as a booster dose for conciseness. The vaccination time ($V_{t}$) for both regimens was assumed to follow the same exponential distribution whose baseline rate parameter $\lambda$ was randomly drawn from a vector of (1/25, 1/50, 1/100, 1/200, 1/300, 1/500) to ensure a well-mixed vaccination elapse duration over the time span. Then, the overall rate parameter of the exponential distribution was set as the product of the baseline rate parameter and the exponential term of the following linear multiplier of $C$ and $C$.

$$\lambda_{V}=\lambda exp(0.2X_{1}+0.5X_{2}-0.5X_{3}-X_{4}+2H)$$

We rounded up the generated vaccination times to the nearest integer number to mimic day-unit vaccination records. To ensure a gap time between the two vaccination regimes, the uptake time of the booster dose was generated at least 20 days after the full doses.

**Second Step: Simulation of 𝐼, 𝑆, and 𝑇**

This simulation incorporated up to two infections per subject: an initial infection and a reinfection. The occurrence times of these infections were generated using exponential distributions with baseline rates of 1/30 and 1/60, respectively. The overall rates were the product of the baseline rates and an exponential term of a linear function of the age group, sex, risk group, and HSB. The coefficients of these variables were assumed to be the same for both infections, given as (𝛽1, 𝛽2, 𝛽3, 𝛽4, 𝛽5) = (0.3, 0.5, −0.2, −1.0, −0.5). The full doses and booster dose were assumed to have constant VE against the initial infection of 50% and 60%, respectively, and against the reinfection, these were 70% and 80%, respectively (eTable 2). eAlgorithm 1 was used to generate data for the constant VE with recurrent infections and time-varying vaccination. A frailty term 𝜃, following a Gamma (5, 0.2) distribution with a mean of 1 and a variance of 0.2, was multiplied on the linear function to add the within-subject correlation. Infection times were rounded up to integer days. An administrative censoring time of 730 days was applied, resulting in censoring rates of about 6% and 40% for the initial infection and reinfection, respectively.

| Infection | Doses | VE |
| --- | --- | --- |
| Initial Infection | 2 | 50% |
| Initial Infection | 3 | 60% |
| Reinfection | 2 | 70% |
| Reinfection | 3 | 80% |

eTable 2 True VE values in the simulation study where VE remains constant over time

We only considered symptomatic infection in our simulation as, by design, the TND study generally only captures symptomatic infection. Therefore, the variable of symptom presentation (𝑆) was always set to 1.

The variable ($D$) indicated testing or not, which was simulated by the following function to ensure that individuals who underwent testing would always have a positive HSB:

$$P\left( D=1 | H, S \right)=S\times(1-\mathrm{expit}(99999-100000\times H))$$

This setting, namely, set that subjects with symptoms and a positive HSB would have a 0.73 probability to test, and subjects with symptoms and 𝐻 = 0 would have a zero probability to test. This guaranteed that all subjects with symptoms who went to testing were with $H=1$.

Finally, we simulated one co-circulating pathogen that could cause symptoms similar to those of COVID-19, prompting subjects with $T=1$ to go for testing even if they were not infected by SARS-CoV-2. These subjects served as controls in the TND study. The infection times for the co-circulating pathogen are simulated using a mixture of ten exponential distributions with different means: (100, 200, 300, 400, 500, 500, 600, 600, 800, 800), respectively.

The results of the simulation studies were summarized using the following metrics to evaluate and compare the performance of the VE models: the averages of VE estimates for the two vaccination regimens against two infections, their empirical bias, average standard errors, empirical standard deviations, coverage of 95% confidence intervals, and mean square errors (MSE).

### 3 Simulation Settings - VE Wanes over Time

VE typically wanes over time in real-world scenarios. To assess the proposed PWP-GT frailty model under more realistic conditions, we simulated infection data in TND studies with VE that wanes as stepwise functions. All other settings were kept the same as those for scenarios with constant VE.

The proposed PWP-GT frailty model can effectively estimate VE that wanes over time in a piecewise manner by incorporating a series of time-varying covariates. In this simulation study analysis, we added time-dependent indicators of vaccination status on days 0, 90, and 180 to the model. The cumulative sums of their coefficients estimate the logarithm of VE at the corresponding vaccination intervals.

### 4 Simulation Detailed Results - VE Remains Constant over Time

The detailed performance metrics of the seven scenarios of study design and model in the simulation were presented in eTable 3-6. These results demonstrate that the PWP-GT frailty model consistently outperforms others across various scenarios where VE remains constant over time.

| Model | Study | True VE | Avg. $\hat{\mathrm{VE}}$ | Emp. Bias | Avg. $SE(\hat{\mathrm{VE}})$ | Emp. $SD(\hat{\mathrm{VE}})$ | 95% CI Cov. | MSE |
| --- | --- | --- | --- | --- | --- | --- | --- | --- |
| PGF | TND | 50% | 49.64% | -0.36% | 1.59% | 1.58% | 94.83% | 0.03% |
| PGR | TND | 50% | 48.26% | -1.74% | 1.58% | 1.58% | 81.83% | 0.06% |
| CL | TND | 50% | 47.01% | -2.99% | 1.84% | 1.78% | 64.67% | 0.12% |
| ULA | TND | 50% | 46.66% | -3.34% | 1.77% | 1.69% | 53.50% | 0.14% |
| UL | TND | 50% | 41.23% | -8.77% | 1.93% | 1.81% | 0.17% | 0.80% |
| PGF | Cohort | 50% | 55.87% | 5.87% | 0.82% | 0.81% | 0.00% | 0.35% |
| PGR | Cohort | 50% | 54.10% | 4.10% | 0.82% | 0.81% | 0.00% | 0.17% |

eTable 3 Detailed simulation results when VE remains constant over time, initial infection, full doses (2 doses)

| Model | Study | True VE | Avg. $\hat{\mathrm{VE}}$ | Emp. Bias | Avg. $SE(\hat{\mathrm{VE}})$ | Emp. $SD(\hat{\mathrm{VE}})$ | 95% CI Cov. | MSE |
| --- | --- | --- | --- | --- | --- | --- | --- | --- |
| PGF | TND | 60% | 59.34% | -0.66% | 1.42% | 1.43% | 92.50% | 0.02% |
| PGR | TND | 60% | 56.79% | -3.21% | 1.42% | 1.41% | 37.17% | 0.12% |
| CL | TND | 60% | 56.32% | -3.68% | 1.67% | 1.55% | 39.33% | 0.16% |
| ULA | TND | 60% | 54.97% | -5.03% | 1.48% | 1.36% | 4.33% | 0.27% |
| UL | TND | 60% | 27.43% | -32.57% | 1.89% | 1.67% | 0.00% | 10.64% |
| PGF | Cohort | 60% | 67.92% | 7.92% | 0.65% | 0.62% | 0.00% | 0.63% |
| PGR | Cohort | 60% | 64.78% | 4.78% | 0.67% | 0.61% | 0.00% | 0.23% |

eTable 4 Detailed simulation results when VE remains constant over time, initial infection, booster dose (3 Doses)

| Model | Study | True VE | Avg. $\hat{\mathrm{VE}}$ | Emp. Bias | Avg. $SE(\hat{\mathrm{VE}})$ | Emp. $SD(\hat{\mathrm{VE}})$ | 95% CI Cov. | MSE |
| --- | --- | --- | --- | --- | --- | --- | --- | --- |
| PGF | TND | 70% | 69.89% | -0.11% | 1.70% | 1.64% | 95.83% | 0.03% |
| PGR | TND | 70% | 69.41% | -0.59% | 1.70% | 1.64% | 96.17% | 0.03% |
| CL | TND | 70% | 67.76% | -2.24% | 1.96% | 1.90% | 80.17% | 0.09% |
| ULA | TND | 70% | 67.30% | -2.70% | 1.94% | 1.90% | 73.50% | 0.11% |
| UL | TND | 70% | 62.53% | -7.47% | 2.21% | 2.12% | 5.00% | 0.60% |
| PGF | Cohort | 70% | 72.11% | 2.11% | 0.70% | 0.75% | 16.83% | 0.05% |
| PGR | Cohort | 70% | 70.86% | 0.86% | 0.71% | 0.77% | 72.67% | 0.01% |

eTable 5 Detailed simulation results when VE remains constant over time, reinfection, full doses (2 doses)

| Model | Study | True VE | Avg. $\hat{\mathrm{VE}}$ | Emp. Bias | Avg. $SE(\hat{\mathrm{VE}})$ | Emp. $SD(\hat{\mathrm{VE}})$ | 95% CI Cov. | MSE |
| --- | --- | --- | --- | --- | --- | --- | --- | --- |
| PGF | TND | 80% | 79.94% | -0.06% | 0.90% | 0.88% | 96.00% | 0.01% |
| PGR | TND | 80% | 79.83% | -0.17% | 0.86% | 0.84% | 96.00% | 0.01% |
| CL | TND | 80% | 77.18% | -2.82% | 1.24% | 1.19% | 35.00% | 0.09% |
| ULA | TND | 80% | 76.30% | -3.70% | 1.18% | 1.16% | 7.83% | 0.15% |
| UL | TND | 80% | 56.94% | -23.06% | 1.76% | 1.64% | 0.00% | 5.35% |
| PGF | Cohort | 80% | 84.01% | 4.01% | 0.33% | 0.35% | 0.00% | 0.16% |
| PGR | Cohort | 80% | 82.64% | 2.64% | 0.34% | 0.35% | 0.00% | 0.07% |

eTable 6 Detailed simulation results when VE remains constant over time, reinfection, booster dose (3 doses)

### 5 Simulation Detailed Results - VE Wanes over Time

The detailed performance metrics of the PWP-GT frailty model in the simulation where VE wanes over time were presented in eTable 7. These results reveal that the proposed model successfully captures and estimates the waning trend of the VEs.

| Infection | Term | True VE | Avg. $\hat{\mathrm{VE}}$ | Emp. Bias | Avg. $SE(\hat{\mathrm{VE}})$ | Emp. $SD(\hat{\mathrm{VE}})$ | 95% CI Cov. | MSE |
| --- | --- | --- | --- | --- | --- | --- | --- | --- |
| Initial | Short | 50% | 49.51% | -0.49% | 1.49% | 1.50% | 95.00% | 0.02% |
| Initial | Medium | 40% | 39.11% | -0.89% | 2.77% | 2.88% | 94.83% | 0.09% |
| Initial | Long | 30% | 28.60% | -1.40% | 4.35% | 4.66% | 92.33% | 0.24% |
| Reinfection | Short | 80% | 79.76% | -0.24% | 1.22% | 1.26% | 93.67% | 0.02% |
| Reinfection | Medium | 70% | 69.72% | -0.28% | 1.55% | 1.55% | 95.67% | 0.02% |
| Reinfection | Long | 60% | 60.63% | 0.63% | 1.76% | 1.69% | 94.17% | 0.03% |

eTable 7 Detailed simulation results when VE wanes over time, full doses (2 doses)

### 6 Simulation Algorithm for Data Generation

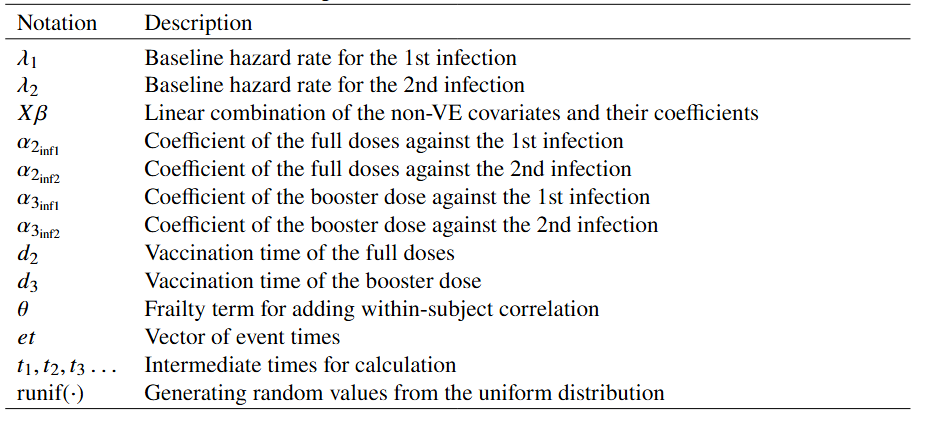

eTable 8 Notation used in the data generation algorithm

eAlgorithm 1. Generate recurrent infection data with time-varying vaccination and within-subject correlation, full and booster doses, constant VE

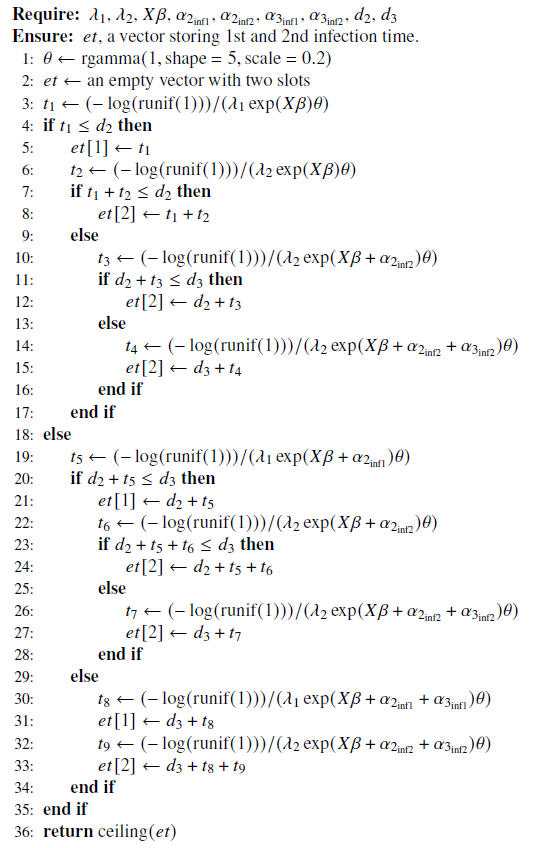

### 7 N3C Data and Its Data Structure

The N3C platform (https://covid.cd2h.org/), sponsored by the US National Institutes of Health (NIH) National Center for Advancing Translational Sciences (NCATS), is an integrated analytical environment that compiles and harmonizes longitudinal electronic health record (EHR) data. It includes data from 84 sites across the United States, covering over 22 million patients. This dataset encompasses individuals who visited testing facilities or healthcare institutes, regardless of whether they tested positive or negative for SARS-CoV-2 or exhibited COVID-19-related symptoms. As one of the largest clinical data collections in the United States for COVID-19 research, it offers a reliable data source for TND studies.

The N3C datasets provide comprehensive details on patient demographics, clinical visits, medical measurements, exposures to medications and devices, clinical conditions, healthcare providers, and observation periods. To ensure the accuracy and reliability of temporal and spatial confounding control, we utilized the most accurate datasets (level-3) in N3C, which include the most transparent timing and location information without random shifting and masking. The N3C database consists of multiple structured data tables. For our analysis, we processed, linked, and combined data from nine primary tables. Below is a brief description of each of these nine tables used in our analysis:

The **Person** table serves as the cornerstone within the entire database, housing comprehensive demographic and clinical details of individual patients. It includes essential variables such as patient ID, date of birth, data partner ID, race, ethnicity, gender, and more, forming a primary table to link to other tables.

The **Location** table comprises data regarding the geographic locations linked with patients and their respective healthcare facilities where data collection occurred. Its variables encompass details such as addresses, states, countries, zip codes, and more, facilitating spatial analysis and contextual understanding within the dataset.

The **Measurement** table records clinical measurements, laboratory results, and observations documented during patient encounters. It has variables such as measurement names, timestamps, and measurement results. This table includes the COVID-19 testing records (PCR or Antigen tests).

The **Drug Exposure** table documents patients’ exposure to various medications and treatments. It provides a comprehensive record of pharmaceutical interventions, including details such as drug names, dosage regimens, administration routes, and duration of exposure. This table includes records for tracking COVID-19 vaccination.

The **Procedure** **Occurrence** table captures a spectrum of medical procedures performed during patient encounters. Key variables include procedure names, timestamps, and associated patient IDs. This table serves as a supplementary resource for the records of COVID-19 vaccination.

The **Manifest** table stores metadata of administrative and data partners. It catalogs the datasets contributed by various healthcare organizations, detailing information such as dataset names, data partner IDs, data types, and data availability timestamps. Key variables include data partner IDs, Common Data Model (CDM) names and versions, and date shift info.

The **Condition** **Occurrence** table has a wide range of medical conditions diagnosed during patient encounters. This table captures essential information such as condition names, timestamps, and associated patient IDs, providing comprehensive insights into the prevalence and management of various health conditions. This table provides information to determine the occurrence of COVID-19-related symptoms.

The **Observation** **Period** table includes the start and end dates of observation periods, patient IDs, and additional relevant metadata critical for longitudinal studies.

The **Concept** **Set** **Members** table organizes and categorizes clinical concepts and their associations within the dataset. This table contains information about the membership of specific clinical concepts within predefined sets or groups, facilitating efficient data management and analysis.

### 8 N3C Data Processing and Model Fitting

In accordance with the data security and privacy requirements of the N3C platform, the data management and analysis for this study were conducted within the N3C Data Enclave. This secure computing platform, based on Palantir Foundry, is designed to streamline data processing while ensuring the security and privacy of individual information. The analysis scripts were written using Apache Spark, Python, and R.

The data processing was divided into two distinct parts: one for testing data and the other for vaccination data. This separation enabled clearly specialized filtering and arrangement of each part, optimizing processing data flows. eFigure 2 provides an overall visual representation of the data processing and model-fitting workflows.

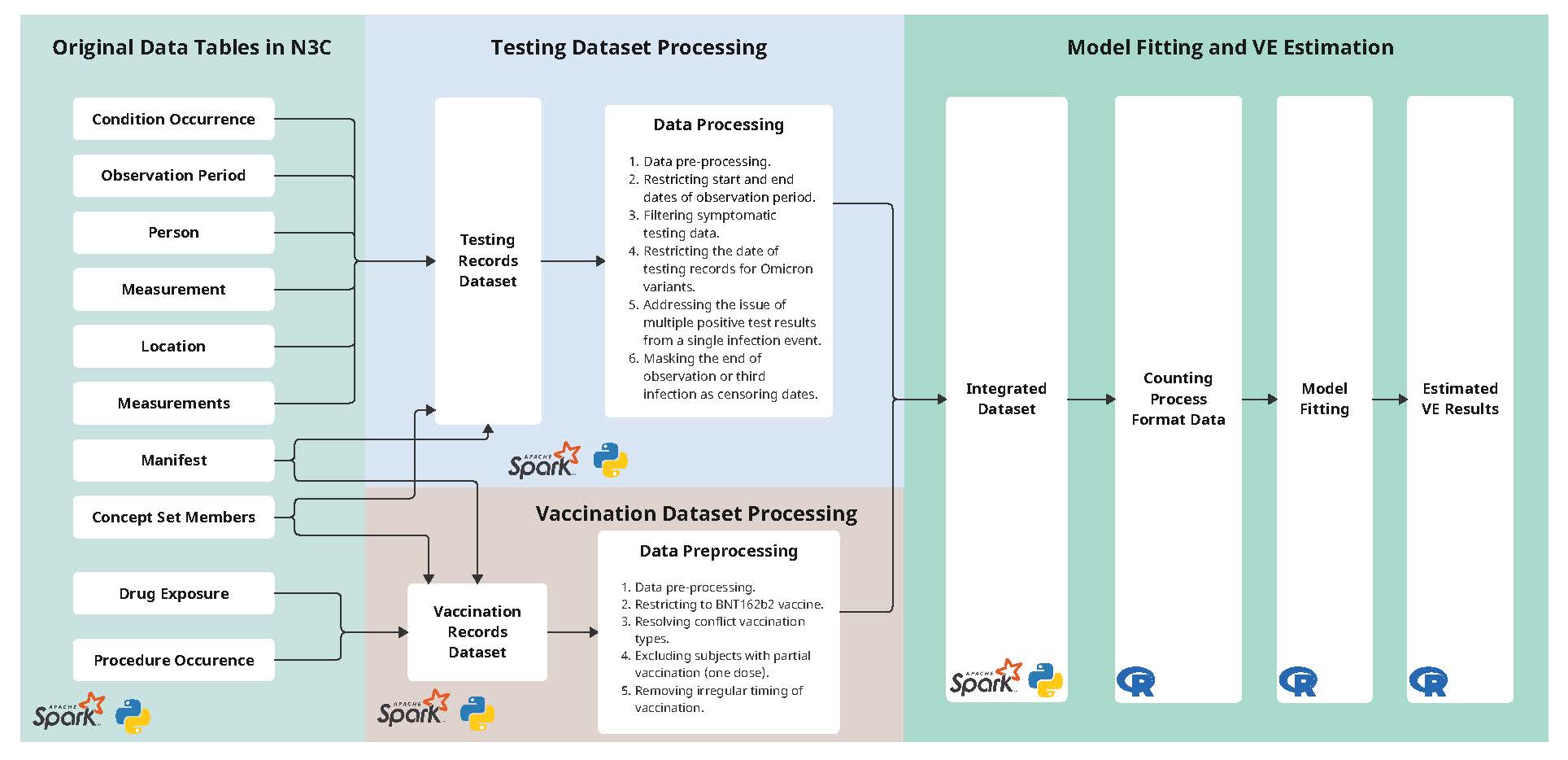

eFigure 2 Flowchart of N3C data processing and analysis.

**Testing Dataset Processing**

Testing data was integrated by the following steps using eight N3C data tables: Condition Occurrence, Observation Period, Person, Measurement, Location, Measurements, Manifest, and Concept Set Members.

First, a series of data pre-processing procedures were done to convert data entries, remove redundant text, and unify types of testing records.

Second, we retained subjects whose observation periods started between January 1, 2000, and January 1, 2020, and ended between January 1, 2023, and April 14, 2024. This filtering helped eliminate anomalies in start or end dates and maintained a focus on individuals with adequately long observation spans.

Third, following the principle of the TND study, we included only COVID-19 testing records for individuals exhibiting COVID-19-related symptoms as defined by the N3C codeset ”CDC covid symptoms UC” (ID: 82402523) within the Concept Set Members table. This codeset includes a total of 565 potential COVID-19-related symptoms, such as sweating, fever, cough with fever, muscle fatigue, and vomiting due to infectious disease. Additionally, a testing record is considered associated with the onset of symptoms only if the testing date is within 7 days before or up to 14 days after the onset of symptoms.

Fourth, we excluded testing records dated before December 1, 2021, when the first U.S. case of COVID-19 attributed to the Omicron variant was reported. This restriction made the initial infection and reinfection in this analysis specifically for the Omicron variants. The prior infection of the variants before Omicron was not considered.

Fifth, we removed any positive COVID-19 test results occurring within a 60-day span following an initial positive test to address the issue of multiple positive test results from a single infection event. we did not account for false positive or negative testing results, given that COVID-19 PCR and antigen tests have relatively high sensitivity (80%) and very high specificity (99%).

Finally, subjects will be censored either at the end of their observation period or at the time of their third infection, as our focus is on VE against the first and second COVID-19 infections.

**Vaccination Dataset Processing**

The vaccination data were joined by four N3C data tables: Drug Exposure, Procedure Occurrence, Manifest, and Concept Set Members. Vaccination records of the Pfizer-BioNTech BNT162b2 vaccine were coded as specific IDs of drug or medical procedures, such as 37003436, 1759206, and 759694, in the Drug Exposure or Procedure Occurrence tables.

First, a series of data pre-processing procedures were done to unify vaccine names, remove incorrect vaccine types, and combine redundant vaccination records.

Second, we excluded vaccination records before December 11, 2020, when the focusing on the Pfizer-BioNTech COVID-19 vaccine (BNT162b2) became available under Emergency Use Authorization (EUA) for individuals aged 16 and older. We also excluded vaccination records whose dates were later than the date of data reporting.

Third, we set vaccination records as missing values when they showed conflicting vaccine types on the same day.

Fourth, we excluded any vaccination records that did not correspond to the Pfizer-BioNTech COVID-19 vaccine.

Fifth, we removed subjects with only partial vaccination (i.e., only the first dose received).

Sixth, a series of exclusion rules were applied to remaining vaccination records with irregular timing:

•	Any doses administered less than 18 days apart were excluded.

•	A gap of more than 42 days between the first and second doses led to exclusion.

•	The gap between the second and third doses being less than five months or more than twelve months resulted in exclusion.

•	Intervals among subsequent booster doses that were more than ten months or less than five months apart led to exclusion.

•	Records where a monovalent vaccine was administered after a bivalent vaccine were excluded.

In the last step, the final dataset in the analysis was obtained by joining the testing dataset with the vaccination dataset by matching the person IDs.

**Model Fitting**

The pre-requisite step to fit the PWP-GT frailty is to prepare the dataset into a counting process format. This was done by the *survival::tmerge* function in R. The gap-time format was prepared by subtracting the previous event time from the time of the following infection events.

The pre-Omicron infection status was treated as a categorical variable 0, 1, >= 2.

The case numbers were treated as time-dependent variables and calculated for each four-month period. Before being incorporated into the model, they were normalized by dividing by their average and then transformed using a logarithmic scale, i.e., log(x/mean(x)) where x represents a vector of four-month case numbers.

To estimate VE, four time-varying covariates were added for full doses and booster doses at 14-180 days and 180+ days, respectively. The model fitting is implemented by the *coxme::coxme* function in R. The frailty term was set with default in *coxme::coxme*. The proportional hazards assumption was checked by graphs of the scaled Schoenfeld residuals output from the *survival::cox.zph* function.

### 9 Distribution of Participants Characteristics in the N3C Data

eFigure 3-6 displayed the distribution of age, location (state), infection time, and vaccination time for the included subjects in the final dataset of this analysis.

Note: all bins in the histogram with counts less than 20 were truncated to 0 due to privacy policy of N3C.

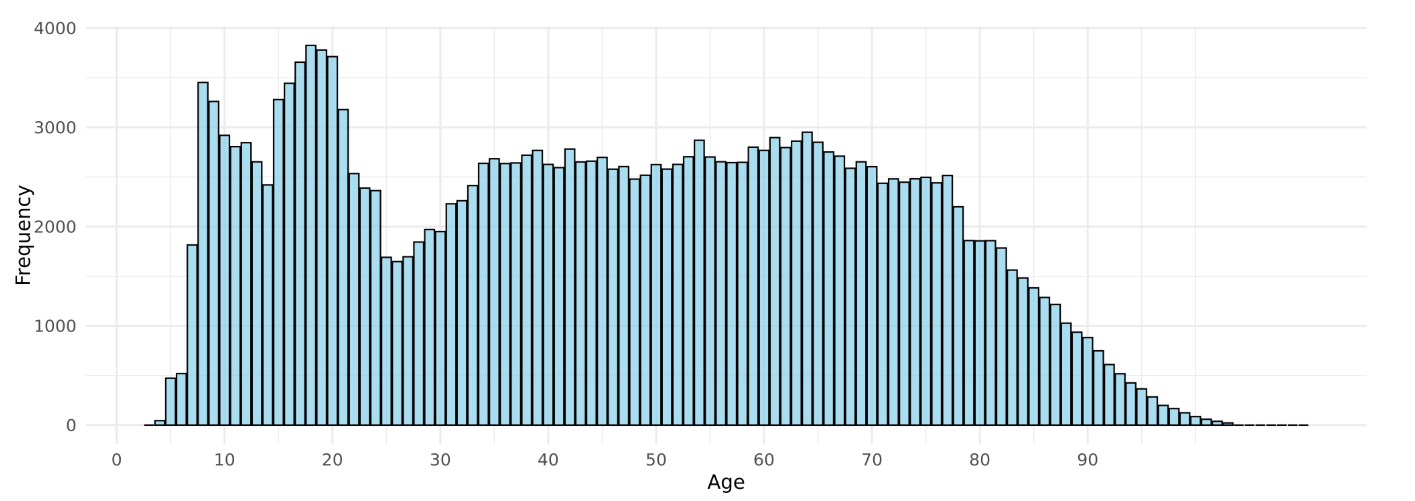

eFigure 3 Histogram of age of the N3C dataset in the analysis.

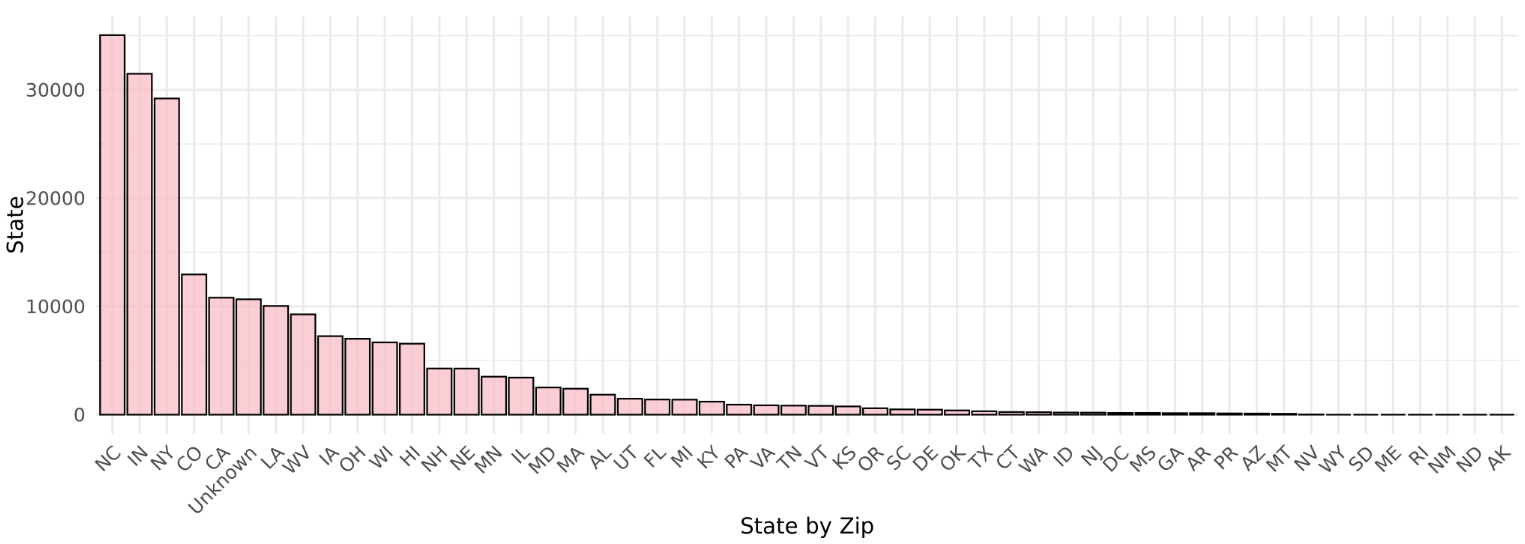

eFigure 4 Barplot of location (state) of the N3C dataset in the analysis.

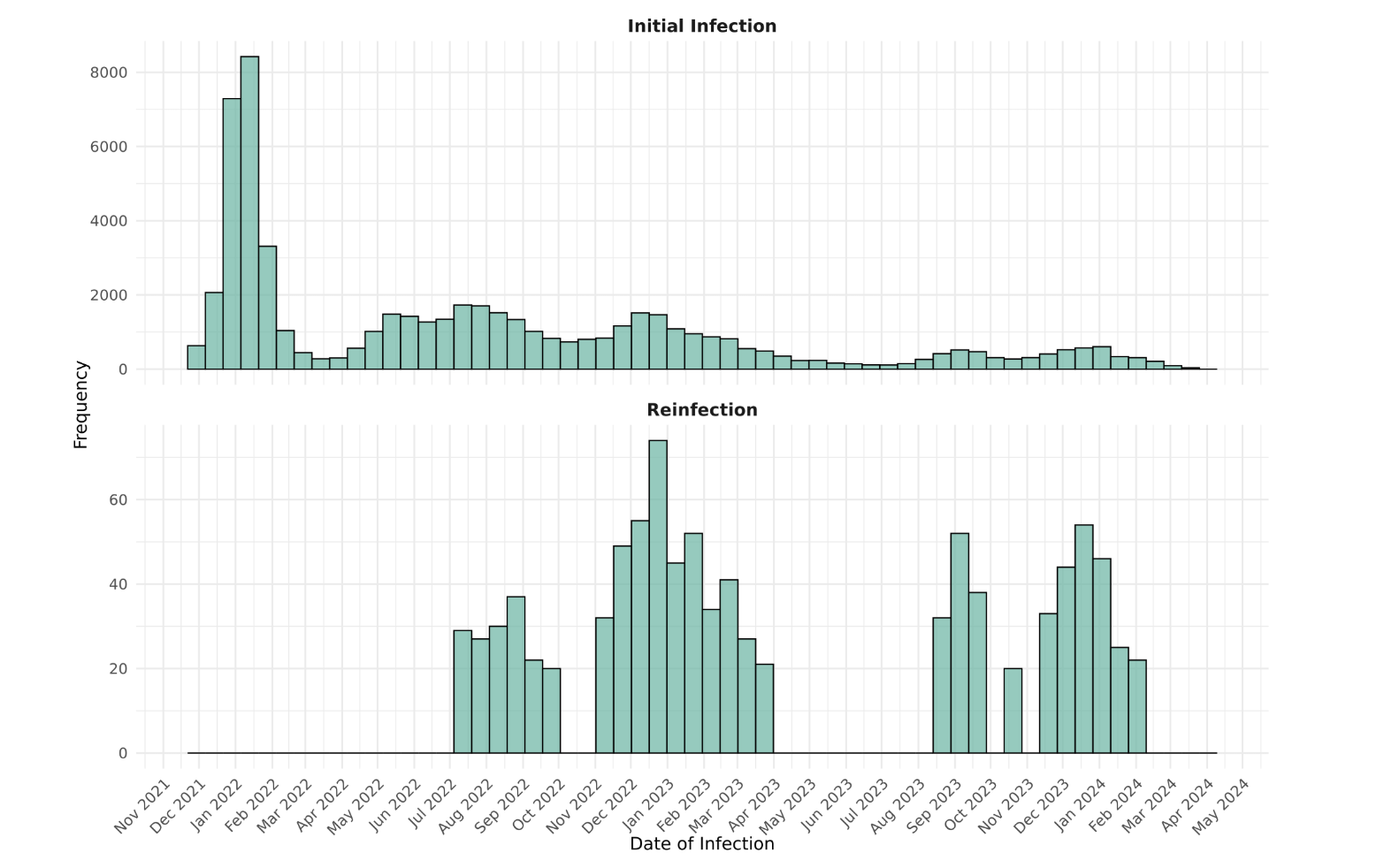

eFigure 5 Histogram of the initial infection and reinfection time of the N3C dataset in the analysis.

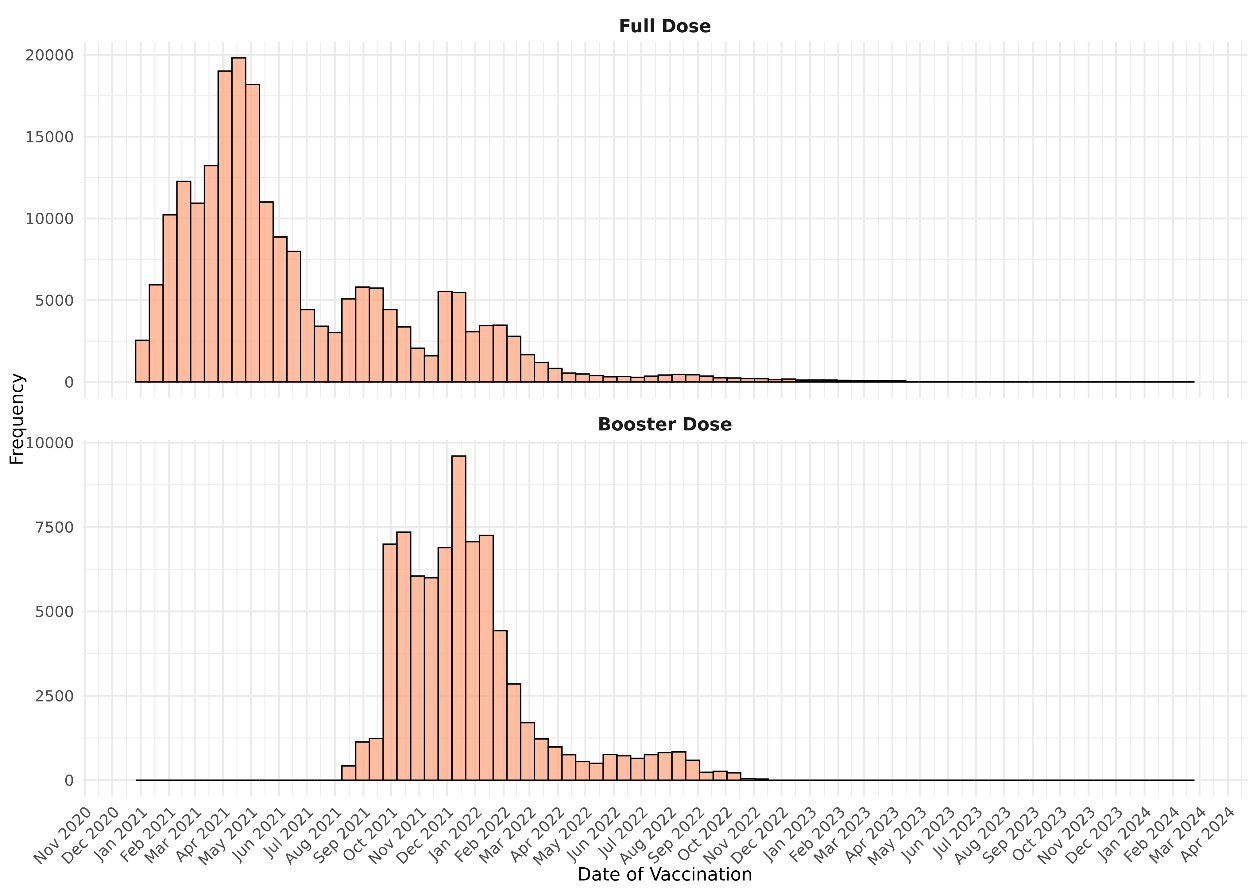

eFigure 6 Histogram of vaccination time of the N3C dataset in the analysis.

### 10 Supplementary Acknowledgement

The N3C Publication committee confirmed that this manuscript msid:2339.618 is in accordance with N3C data use and attribution policies; however, this content is solely the responsibility of the authors and does not necessarily represent the official views of the National Institutes of Health or the N3C program.

We gratefully acknowledge the following core contributors to N3C: Adam B. Wilcox, Adam M. Lee, Alexis Graves, Alfred (Jerrod) Anzalone, Amin Manna, Amit Saha, Amy Olex, Andrea Zhou, Andrew E. Williams, Andrew M. Southerland, Andrew T. Girvin, Anita Walden, Anjali Sharathkumar, Benjamin Amor, Benjamin Bates, Brian Hendricks, Brijesh Patel, G. Caleb Alexander, Carolyn T. Bramante, Cavin Ward-Caviness, Charisse Madlock-Brown, Christine Suver, Christopher G. Chute, Christopher Dillon, Chunlei Wu, Clare Schmitt, Cliff Takemoto, Dan Housman, Davera Gabriel, David A. Eichmann, Diego Mazzotti, Donald E. Brown, Eilis Boudreau, Elaine L. Hill, Emily Carlson Marti, Emily R. Pfaff, Evan French, Farrukh M Koraishy, Federico Mariona, Fred Prior, George Sokos, Greg Martin, Harold P. Lehmann, Heidi Spratt, Hemalkumar B. Mehta, J.W. Awori Hayanga, Jami Pincavitch, Jaylyn Clark, Jeremy Richard Harper, Jessica Yasmine Islam, Jin Ge, Joel Gagnier, Johanna J. Loomba, John B. Buse, Jomol Mathew, Joni L. Rutter, Julie A. McMurry, Justin Guinney, Justin Starren, Karen Crowley, Katie Rebecca Bradwell, Kellie M. Walters, Ken Wilkins, Kenneth R. Gersing, Kenrick Cato, Kimberly Murray, Kristin Kostka, Lavance Northington, Lee Pyles, Lesley Cottrell, Lili M. Portilla, Mariam Deacy, Mark M. Bissell, Marshall Clark, Mary Emmett, Matvey B. Palchuk, Melissa A. Haendel, Meredith Adams, Meredith Temple-O'Connor, Michael G. Kurilla, Michele Morris, Nasia Safdar, Nicole Garbarini, Noha Sharafeldin, Ofer Sadan, Patricia A. Francis, Penny Wung Burgoon, Philip R.O. Payne, Randeep Jawa, Rebecca Erwin-Cohen, Rena C. Patel, Richard A. Moffitt, Richard L. Zhu, Rishikesan Kamaleswaran, Robert Hurley, Robert T. Miller, Saiju Pyarajan, Sam G. Michael, Samuel Bozzette, Sandeep K. Mallipattu, Satyanarayana Vedula, Scott Chapman, Shawn T. O'Neil, Soko Setoguchi, Stephanie S. Hong, Steven G. Johnson, Tellen D. Bennett, Tiffany J. Callahan, Umit Topaloglu, Valery Gordon, Vignesh Subbian, Warren A. Kibbe, Wenndy Hernandez, Will Beasley, Will Cooper, William Hillegass, Xiaohan Tanner Zhang.

The following institutions whose data is released or pending:

Available: Advocate Health Care Network — UL1TR002389: The Institute for Translational Medicine (ITM) • Aurora Health Care Inc — UL1TR002373: Wisconsin Network For Health Research • Boston University Medical Campus — UL1TR001430: Boston University Clinical and Translational Science Institute • Brown University — U54GM115677: Advance Clinical Translational Research (Advance-CTR) • Carilion Clinic — UL1TR003015: iTHRIV Integrated Translational health Research Institute of Virginia • Case Western Reserve University — UL1TR002548: The Clinical & Translational Science Collaborative of Cleveland (CTSC) • Charleston Area Medical Center — U54GM104942: West Virginia Clinical and Translational Science Institute (WVCTSI) • Children’s Hospital Colorado — UL1TR002535: Colorado Clinical and Translational Sciences Institute • Columbia University Irving Medical Center — UL1TR001873: Irving Institute for Clinical and Translational Research • Dartmouth College — None (Voluntary) Duke University — UL1TR002553: Duke Clinical and Translational Science Institute • George Washington Children’s Research Institute — UL1TR001876: Clinical and Translational Science Institute at Children’s National (CTSA-CN) • George Washington University — UL1TR001876: Clinical and Translational Science Institute at Children’s National (CTSA-CN) • Harvard Medical School — UL1TR002541: Harvard Catalyst • Indiana University School of Medicine — UL1TR002529: Indiana Clinical and Translational Science Institute • Johns Hopkins University — UL1TR003098: Johns Hopkins Institute for Clinical and Translational Research • Louisiana Public Health Institute — None (Voluntary) • Loyola Medicine — Loyola University Medical Center • Loyola University Medical Center — UL1TR002389: The Institute for Translational Medicine (ITM) • Maine Medical Center — U54GM115516: Northern New England Clinical & Translational Research (NNE-CTR) Network • Mary Hitchcock Memorial Hospital & Dartmouth Hitchcock Clinic — None (Voluntary) • Massachusetts General Brigham — UL1TR002541: Harvard Catalyst • Mayo Clinic Rochester — UL1TR002377: Mayo Clinic Center for Clinical and Translational Science (CCaTS) • Medical University of South Carolina — UL1TR001450: South Carolina Clinical & Translational Research Institute (SCTR) • MITRE Corporation — None (Voluntary) • Montefiore Medical Center — UL1TR002556: Institute for Clinical and Translational Research at Einstein and Montefiore • Nemours — U54GM104941: Delaware CTR ACCEL Program • NorthShore University HealthSystem — UL1TR002389: The Institute for Translational Medicine (ITM) • Northwestern University at Chicago — UL1TR001422: Northwestern University Clinical and Translational Science Institute (NUCATS) • OCHIN — INV-018455: Bill and Melinda Gates Foundation grant to Sage Bionetworks • Oregon Health & Science University — UL1TR002369: Oregon Clinical and Translational Research Institute • Penn State Health Milton S. Hershey Medical Center — UL1TR002014: Penn State Clinical and Translational Science Institute • Rush University Medical Center — UL1TR002389: The Institute for Translational Medicine (ITM) • Rutgers, The State University of New Jersey — UL1TR003017: New Jersey Alliance for Clinical and Translational Science • Stony Brook University — U24TR002306 • The Alliance at the University of Puerto Rico, Medical Sciences Campus — U54GM133807: Hispanic Alliance for Clinical and Translational Research (The Alliance) • The Ohio State University — UL1TR002733: Center for Clinical and Translational Science • The State University of New York at Buffalo — UL1TR001412: Clinical and Translational Science Institute • The University of Chicago — UL1TR002389: The Institute for Translational Medicine (ITM) • The University of Iowa — UL1TR002537: Institute for Clinical and Translational Science • The University of Miami Leonard M. Miller School of Medicine — UL1TR002736: University of Miami Clinical and Translational Science Institute • The University of Michigan at Ann Arbor — UL1TR002240: Michigan Institute for Clinical and Health Research • The University of Texas Health Science Center at Houston — UL1TR003167: Center for Clinical and Translational Sciences (CCTS) • The University of Texas Medical Branch at Galveston — UL1TR001439: The Institute for Translational Sciences • The University of Utah — UL1TR002538: Uhealth Center for Clinical and Translational Science • Tufts Medical Center — UL1TR002544: Tufts Clinical and Translational Science Institute • Tulane University — UL1TR003096: Center for Clinical and Translational Science • The Queens Medical Center — None (Voluntary) • University Medical Center New Orleans — U54GM104940: Louisiana Clinical and Translational Science (LA CaTS) Center • University of Alabama at Birmingham — UL1TR003096: Center for Clinical and Translational Science • University of Arkansas for Medical Sciences — UL1TR003107: UAMS Translational Research Institute • University of Cincinnati — UL1TR001425: Center for Clinical and Translational Science and Training • University of Colorado Denver, Anschutz Medical Campus — UL1TR002535: Colorado Clinical and Translational Sciences Institute • University of Illinois at Chicago — UL1TR002003: UIC Center for Clinical and Translational Science • University of Kansas Medical Center — UL1TR002366: Frontiers: University of Kansas Clinical and Translational Science Institute • University of Kentucky — UL1TR001998: UK Center for Clinical and Translational Science • University of Massachusetts Medical School Worcester — UL1TR001453: The UMass Center for Clinical and Translational Science (UMCCTS) • University Medical Center of Southern Nevada — None (voluntary) • University of Minnesota — UL1TR002494: Clinical and Translational Science Institute • University of Mississippi Medical Center — U54GM115428: Mississippi Center for Clinical and Translational Research (CCTR) • University of Nebraska Medical Center — U54GM115458: Great Plains IDeA-Clinical & Translational Research • University of North Carolina at Chapel Hill — UL1TR002489: North Carolina Translational and Clinical Science Institute • University of Oklahoma Health Sciences Center — U54GM104938: Oklahoma Clinical and Translational Science Institute (OCTSI) • University of Pittsburgh — UL1TR001857: The Clinical and Translational Science Institute (CTSI) • University of Pennsylvania — UL1TR001878: Institute for Translational Medicine and Therapeutics • University of Rochester — UL1TR002001: UR Clinical & Translational Science Institute • University of Southern California — UL1TR001855: The Southern California Clinical and Translational Science Institute (SC CTSI) • University of Vermont — U54GM115516: Northern New England Clinical & Translational Research (NNE-CTR) Network • University of Virginia — UL1TR003015: iTHRIV Integrated Translational health Research Institute of Virginia • University of Washington — UL1TR002319: Institute of Translational Health Sciences • University of Wisconsin-Madison — UL1TR002373: UW Institute for Clinical and Translational Research • Vanderbilt University Medical Center — UL1TR002243: Vanderbilt Institute for Clinical and Translational Research • Virginia Commonwealth University — UL1TR002649: C. Kenneth and Dianne Wright Center for Clinical and Translational Research • Wake Forest University Health Sciences — UL1TR001420: Wake Forest Clinical and Translational Science Institute • Washington University in St. Louis — UL1TR002345: Institute of Clinical and Translational Sciences • Weill Medical College of Cornell University — UL1TR002384: Weill Cornell Medicine Clinical and Translational Science Center • West Virginia University — U54GM104942: West Virginia Clinical and Translational Science Institute (WVCTSI)  Submitted: Icahn School of Medicine at Mount Sinai — UL1TR001433: ConduITS Institute for Translational Sciences • The University of Texas Health Science Center at Tyler — UL1TR003167: Center for Clinical and Translational Sciences (CCTS) • University of California, Davis — UL1TR001860: UCDavis Health Clinical and Translational Science Center • University of California, Irvine — UL1TR001414: The UC Irvine Institute for Clinical and Translational Science (ICTS) • University of California, Los Angeles — UL1TR001881: UCLA Clinical Translational Science Institute • University of California, San Diego — UL1TR001442: Altman Clinical and Translational Research Institute • University of California, San Francisco — UL1TR001872: UCSF Clinical and Translational Science Institute  NYU Langone Health Clinical Science Core, Data Resource Core, and PASC Biorepository Core — OTA-21-015A: Post-Acute Sequelae of SARS-CoV-2 Infection Initiative (RECOVER)  Pending: Arkansas Children’s Hospital — UL1TR003107: UAMS Translational Research Institute • Baylor College of Medicine — None (Voluntary) • Children’s Hospital of Philadelphia — UL1TR001878: Institute for Translational Medicine and Therapeutics • Cincinnati Children’s Hospital Medical Center — UL1TR001425: Center for Clinical and Translational Science and Training • Emory University — UL1TR002378: Georgia Clinical and Translational Science Alliance • HonorHealth — None (Voluntary) • Loyola University Chicago — UL1TR002389: The Institute for Translational Medicine (ITM) • Medical College of Wisconsin — UL1TR001436: Clinical and Translational Science Institute of Southeast Wisconsin • MedStar Health Research Institute — None (Voluntary) • Georgetown University — UL1TR001409: The Georgetown-Howard Universities Center for Clinical and Translational Science (GHUCCTS) • MetroHealth — None (Voluntary) • Montana State University — U54GM115371: American Indian/Alaska Native CTR • NYU Langone Medical Center — UL1TR001445: Langone Health’s Clinical and Translational Science Institute • Ochsner Medical Center — U54GM104940: Louisiana Clinical and Translational Science (LA CaTS) Center • Regenstrief Institute — UL1TR002529: Indiana Clinical and Translational Science Institute • Sanford Research — None (Voluntary) • Stanford University — UL1TR003142: Spectrum: The Stanford Center for Clinical and Translational Research and Education • The Rockefeller University — UL1TR001866: Center for Clinical and Translational Science • The Scripps Research Institute — UL1TR002550: Scripps Research Translational Institute • University of Florida — UL1TR001427: UF Clinical and Translational Science Institute • University of New Mexico Health Sciences Center — UL1TR001449: University of New Mexico Clinical and Translational Science Center • University of Texas Health Science Center at San Antonio — UL1TR002645: Institute for Integration of Medicine and Science • Yale New Haven Hospital — UL1TR001863: Yale Center for Clinical Investigation
